# Structural Brain Biomarkers for Identifying Progressive Amnestic Mild Cognitive Impairment

**DOI:** 10.64898/2026.07.27.26359003

**Authors:** Nikita Cherkasov, Aleksandr Tomyshev, Ekaterina Abdullina, Anastasiya Dudina, Yana Fedorova, Elena Ponomareva, Natalia Selezneva, Olga Bozhko, Svetlana Gavrilova, Igor Kolykhalov, Irina Lebedeva

## Abstract

**Objective:** To identify the structural features of the brain in patients with amnestic mild cognitive impairment (aMCI) compared to healthy individuals, as well as to detect structural markers of progression to dementia in aMCI subgroups with different clinical outcomes.

**Materials and Methods:** 41 patients with aMCI (mean age 71.5±9.5 years, 33 females) who underwent clinical assessment during 1 year follow-up, and 38 healthy controls (mean age 63.7±11.7 years, 28 females) were included. MRI scans were acquired, and the cortical thickness in both hemispheres, as well as the volumes of subcortical structures and hippocampal subfields, were analyzed.

**Results:** Patients with aMCI as a whole, as well as aMCI subgroups with different clinical outcomes, showed significantly smaller bilateral hippocampal volumes and volumes of several hippocampal subfields than healthy controls. In addition, patients who were subsequently diagnosed with Alzheimer’s disease dementia exhibited lower cortical gray matter thickness and smaller bilateral amygdala volumes than the healthy controls. Several subcortical structures and hippocampal subregions remained preserved in patients with aMCI, including those with unfavorable clinical outcomes.

**Conclusion:** The study revealed structural brain abnormalities in aMCI, characterized by reduced hippocampal and hippocampal subfield volumes, as well as amygdala volume reduction, while other subcortical structures were relatively preserved. Furthermore, reduced cortical gray matter thickness may serve as a potential prognostic marker for conversion to dementia in patients with aMCI.

## Introduction

Amnestic mild cognitive impairment (aMCI) is widely regarded as a potential prodromal stage of Alzheimer’s disease (AD). In recent years, a large number of MRI studies have been conducted with two main objectives: first, to identify structural and functional brain characteristics specific to aMCI and second, to detect neuroimaging markers capable of reliably predicting conversion from aMCI to dementia. It has been demonstrated that aMCI is associated with numerous aberrant structural brain features, including gray matter atrophy in the hippocampus, parahippocampal gyrus, amygdala, thalamus, and precuneus ^1–5^. Smirnova et al. reported significant between-group differences between the non-amnestic and amnestic MCI subtypes in the volumes of the anterior corpus callosum, right caudate nucleus, left cerebellar hemispheric cortex, posterior corpus callosum, and left thalamus ^6^. Atrophy of the anterior corpus callosum and right caudate nucleus has been proposed as a biomarker of unfavorable prognosis.

The assessment of hippocampal structural characteristics is of particular interest to researchers, as this structure plays a central role in memory function and is among the earliest and most severely affected by neurodegenerative processes, showing progressive morphological changes as aMCI advances to AD. Risacher et al. argued that hippocampal volume reduction represents the best and most reliable MRI marker for imminent conversion from MCI to AD ^7^. In particular, greater volume loss was observed in the left hippocampus, predominantly involving the CA1 subfield and dentate gyrus, as well as both the head and tail of the hippocampus ^8^. The inclusion of hippocampal subfield analysis in MRI data has enabled the detection of significant differences between patients with aMCI and controls in gray matter volumes of the subiculum, presubiculum, molecular layer of the hippocampus, CA1, and CA4 ^9^. In another study by Stulov et al., a model incorporating the left subiculum volume, right entorhinal cortex thickness, and volume fraction of hypointense (vascular) lesions achieved 85% accuracy in distinguishing the aMCI subgroup (considered by the authors to be at the prodromal stage of AD dementia) from the subgroup with subcortical vascular etiology of MCI ^10^.

A considerably smaller number of studies have examined morphological brain features in aMCI subgroups with different clinical outcomes. For example, in a multicenter study by Brueggen et al., a reduced right hippocampal volume was identified as a predictor of conversion to dementia in patients with MCI ^11^. In a study by Nesteruk et al., the best predictive performance for progression from MCI to Alzheimer’s disease dementia was achieved by a model that included volumes of the left hippocampus and left entorhinal cortex (although the sensitivity of the method remained relatively low at 59%) ^12^. In their review, Drago et al. identified markers capable of determining disease trajectory, describing structural patterns characteristic of patients with progressive MCI versus those with stable MCI ^13^. The so-called “Class A markers”, which possess a high degree of validity, include characteristics reported in several longitudinal studies. Among these, there was a greater rate of cortical volume loss, ventricular enlargement, and hippocampal volume reduction in patients with the progressive form of MCI. The majority of the studies included in this review ^13^ comprised samples of patients with aMCI.

These structural neuroimaging findings are complemented by recent clinical-biological research demonstrating that neuroinflammation serves as a key pathogenetic driver of neurodegeneration and aMCI progression to dementia. Ponomareva et al. ^14^ established that baseline serum levels of pro-inflammatory cytokines and immune-response indicators reliably predict three-year outcomes. Specifically, elevated IL-1β, IL-6, IL-2, and TNFα, together with reduced enzymatic activity of leukocyte elastase, significantly differentiated patients with unfavorable outcomes from those with stable or improved cognition during 3-year follow-up. These blood-based neuroimmune markers offer a prognostic tool that likely reflect the underlying mechanisms of the hippocampal atrophy and cortical volume loss observed on MRI, thereby enhancing multimodal risk stratification beyond structural imaging alone.

Nevertheless, the use of morphometric measures for prognostic purposes, specifically to assess the likelihood of aMCI conversion to dementia, has inherent limitations. In a Cochrane systematic review conducted by Lombardi et al., relatively low sensitivity and specificity were reported for such models ^15^. However, models based on hippocampal volume have demonstrated the highest reliability, outperforming other models, such as those using medial temporal lobe atrophy or lateral ventricular volume ^15,16^. Furthermore, the inclusion of hippocampal subfield morphometric parameters rather than total hippocampal volume alone increases the accuracy of these predictive models ^17^. Although structural MRI cannot serve as the sole criterion for evaluating dementia risk, the incorporation of morphometric parameters into predictive models can form an important component of multidimensional prognostic assessment in patients with MCI.

In light of the above, further investigation of the structural brain characteristics in MCI and aMCI, along with identification of potential prognostic markers of conversion to dementia, remains essential. Thus, the aim of the present study was to identify structural brain features associated with aMCI and to determine potential neuroimaging markers capable of predicting the conversion from aMCI to AD dementia through the analysis of structural MRI data.

## Materials and Methods

***The inclusion criteria*** for the clinical group were as follows: age ≥ 50 years; amnestic mild cognitive impairment verified according to the diagnostic criteria proposed by Winblad et al. ^18^.

***Exclusion criteria*** for both the clinical and control groups included: a diagnosis of dementia of any etiology, neurological disease (congenital and/or acquired metabolic encephalopathies, toxic and drug-induced encephalopathies, Parkinson’s disease, history of stroke, epilepsy, infectious diseases, demyelinating and hereditary degenerative diseases of the central nervous system), neoplastic and/or traumatic brain lesions, systemic diseases, malignant extracerebral tumors, mental disorders and delirium, alcohol and/or substance abuse, ACE inhibitors and/or memantine medication, and contraindications to MRI. An additional exclusion criterion for the control group was the presence of possible cognitive impairment as indicated by a Montreal Cognitive Assessment screening score < 26 points.

### Participants and Study Design

In this longitudinal observational cohort study with baseline MRI and 1-year clinical follow-up the total sample consisted of 79 participants, including 41 patients with aMCI (mean age 71.5±9.5 years, 33 females) eligible for follow-up and 38 mentally healthy controls (mean age 63.7±11.7 years, 28 females). All patients were retired at the time of inclusion. All participants were right-handed.

Participants were recruited consecutively from the outpatient clinic between 2024 and 2025. Baseline assessments occurred in 2024; follow-up visits continued in 2025. The final sample size was determined by the number of consecutive eligible patients with aMCI who consented and completed the 1-year follow-up during the study period. Potential selection bias was addressed by consecutive recruitment of eligible patients and by age- and sex-matching for between-group comparisons. Confounding by age and sex was controlled statistically (see Statistical analysis).

All individuals underwent a comprehensive clinical evaluation by a psychiatrist specializing in cognitive disorders at the time of enrolment. This assessment included a clinical interview with the participants and their study partners, medical history, evaluation of mental and neurological status, and cognitive functioning. The Montreal Cognitive Assessment (MoCA) ^19^ was used as a screening tool to objectively detect possible cognitive impairment. Patients with identified cognitive impairment during screening were referred to complete a battery of neuropsychological tests. Brain MRI was performed the same day or the following day. Patients were further advised to undergo regular follow-up visits every 6 months, during which their clinical status was reassessed, and cognitive evaluation was readministered. The diagnosis of dementia in Alzheimer’s disease was established according to the International Classification of Diseases, 10th Revision (ICD-10) criteria.

All patients provided written informed consent to participate in this observational study. This non-interventional observational study was performed in accordance with the principles of the Helsinki Declaration (1964) and its later amendments (1975–2013) and was approved by the Local Ethics Committee of the Federal State Budgetary Scientific Institution “Russian Mental Health Research Centre” (Protocol No. 920 25-Dec-2023) and was conducted in 2024-2025.

### Comparison of Demographic and Clinical Characteristics Between Overall aMCI Group and Controls

The aMCI patient group as a whole and the healthy control group did not differ significantly in sex distribution (χ² = 0.2, p = 0.65) but showed a significant difference in age (71.5 vs. 63.7 years, t(77) = 3.3, p = 0.0017, Cohen’s d = 0.73). This age difference did not allow for complete exclusion of the potential influence of age on the analysis results. Therefore, the maximum possible number of participants from both groups were selected using pairwise age- and sex-matching. As a result, between-group analyses were performed on a subsample consisting of 28 patients with aMCI (mean age 69.1±9.1 years, 5 males, 23 females) and 27 healthy controls (mean age 69.0±9.3 years, 5 males, 22 females) (Table 1).

**Table 1.** Demographic and Psychometric Characteristics of the Comparison Groups.

| Characteristic | Overall groups |  | Matched groups |  | Clinical outcome subgroups |  |
| --- | --- | --- | --- | --- | --- | --- |
|  | aMCI<br>(n=41) | Control<br>(n=38) | aMCI<br>(n=28) | Control<br>(n=27) | Stable<br>aMCI<br>(n=17) | aMCI-AD<br>(n=16) |
| **Number of participants** | 41 | 38 | 28 | 27 | 17 | 16 |
| **Age, years** | 71.5±9.5 | 63.7±11.7 | 69.1±9.1 | 69.0±9.3 | 71.0±10.8 | 74.4±7.6 |
| **Sex, M/F** | 8/33 | 10/28 | 5/23 | 5/22 | 4/13 | 3/13 |
| **MoCA score at MRI** | 23.4±2.8 | 29.0±1.8 | 23.6±3.0 | 28.6±2.0 | 23.3±2.5 | 22.9±2.8 |

Note: Values are presented as means±standard deviations. aMCI = amnesic mild cognitive impairment; aMCI-AD = aMCI converters to Alzheimer’s disease dementia; MoCA = Montreal Cognitive Assessment.

### Longitudinal subgroups

All 41 patients with aMCI included in the present analyses completed the scheduled 1- year follow-up assessment. Based on the follow-up outcomes, three subgroups of patients with aMCI were identified in the total sample: 17 patients with no clinically significant progression (hereinafter referred to as “stable” subgroup; mean age 71.0±10.8 years, 13 females, baseline MoCA score 23.3±2.5); 5 patients with cognitive decline that did not meet the criteria for dementia at the time of re-examination (mean age 72.1±6.0 years, 5 females, baseline MoCA score 24.4±2.6; cognitive decline was defined as a decrease of more than 2 points on the MoCA between baseline and follow-up assessments); and 16 patients who were diagnosed with dementia in AD at follow-up (hereinafter referred to as “aMCI-AD”; mean age 74.4±7.6 years, 13 females, baseline MoCA score 22.9±2.8). Three patients were subsequently excluded from subgroup analyses because they converted to non-Alzheimer’s dementia. Analyses were therefore restricted to participants with complete baseline MRI and 1-year clinical outcome data. The group with cognitive decline was not included in the analysis owing to the small sample size. Patients with stable aMCI and those who subsequently progressed to dementia did not differ significantly in the baseline composite MoCA score (U = 119.5, p = 0.557).

For these two subgroups, patients with stable aMCI (n = 17) and patients who were subsequently diagnosed with AD dementia (n = 16), a common subgroup of mentally healthy controls (n = 24) were also selected. This control subgroup did not differ significantly in sex or age between longitudinal aMCI groups. Pairwise between-group comparisons were performed between the three subgroups.

### Magnetic Resonance Imaging and Image Processing

MRI examinations were performed using a 3T Philips Ingenia scanner (The Netherlands). T1-weighted images were acquired using a turbo field echo (TFE) sequence with the following parameters: TR = 8 ms, TE = 4 ms, flip angle = 8°, voxel size = 0.98 × 0.98 × 1.0 mm, 170 slices, and interslice gap = 0 mm.

T1-weighted images were processed using FreeSurfer software package (version 6.0.0) to obtain detailed anatomical reconstructions for each participant. The FreeSurfer pipeline includes correction for magnetic field inhomogeneity, removal of non-brain tissue, and automated anatomical labeling (e.g., thalamus, hippocampus, cerebral ventricles, etc.) for each voxel ^20^. Cortical surface models were then reconstructed, and cortical thickness measurements were derived using the algorithms described by Fischl et al. ^21^. As a result, mean cortical thickness values (in millimeters) were obtained for both hemispheres, along with gray matter volume measurements (in cubic millimeters) for seven subcortical structures (thalamus, caudate nucleus, putamen, globus pallidus, hippocampus, amygdala, and nucleus accumbens) in each hemisphere (Figure 1), according to the built-in FreeSurfer 6.0.0 atlas.

**Figure 1.**
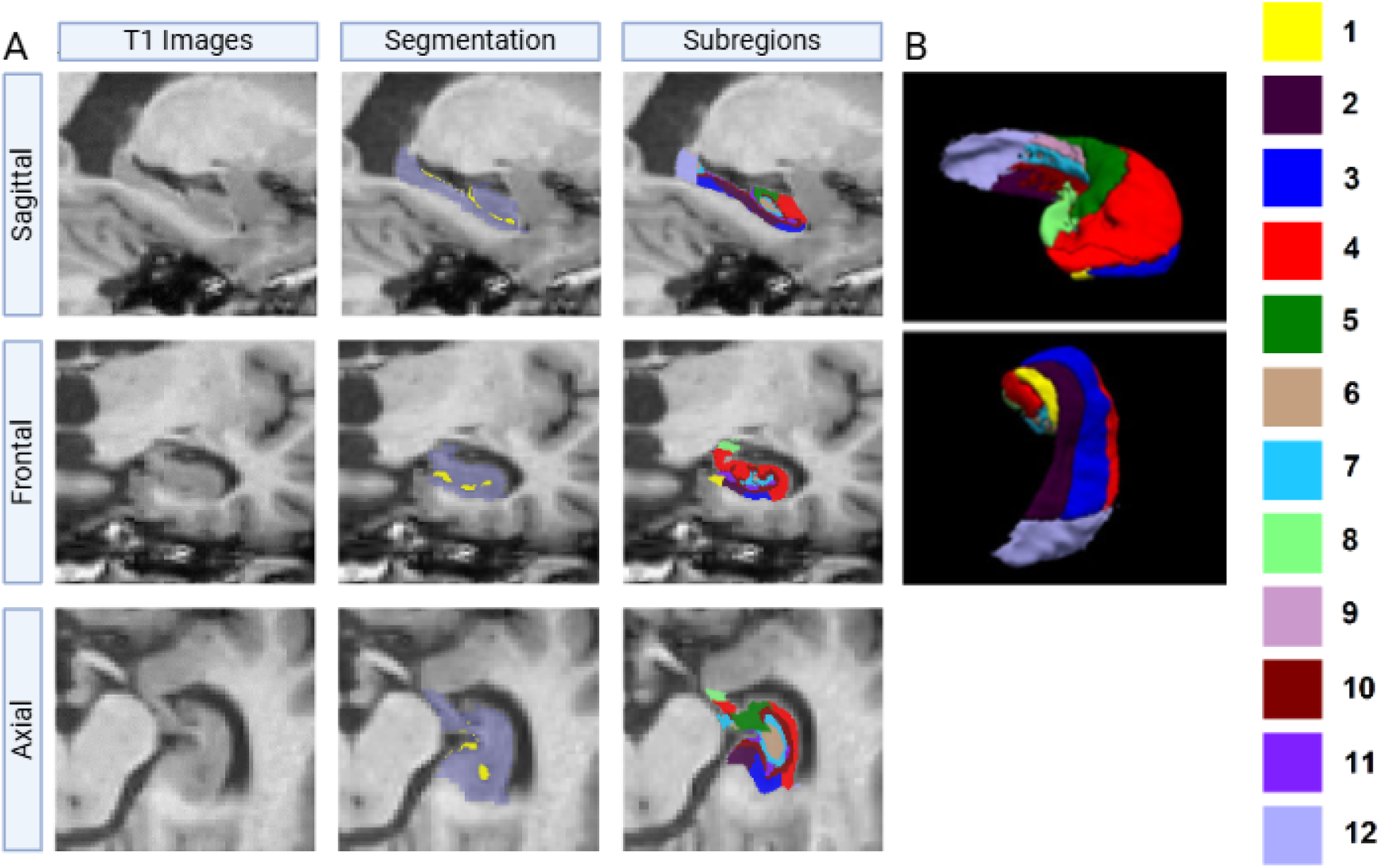
A: Sagittal, frontal and axial slices of the original T1-weighted image (left row), with segmentation of the entire hippocampus (central row) and with segmentation of 12 hippocampal subregions (right row) of a subject with aMCI according to the atlas from Fischl B. ^21^ The names of the subregions according to the color codes are indicated on the right: 1- Parasubiculum; 2- Presubiculum; 3 - Subiculum; subregions 4 - CA1, 5 - CA2/3, 6 - CA4; 7 - GC-DG (Granule Cell Layer of the Dentate Gyrus); 8 - HATA (Hippocampus-Amygdala Transition Area); 9 - Fimbria, 10 - Molecular layer; 11 - Hippocampal fissure; 12 - Hippocampal tail. B: Three-dimensional renderings of the hippocampus segmented using Free Surfer 6.0.0 and hippocampal subregions, modified from Fischl B. ^21^

In addition, 12 hippocampal subregions were reconstructed bilaterally for each participant using previously described algorithms ^22^ (Figure 2). Quality control of the hippocampal and hippocampal subregion segmentations was performed in accordance with a procedure outlined previously ^23^.

**Figure 2.**
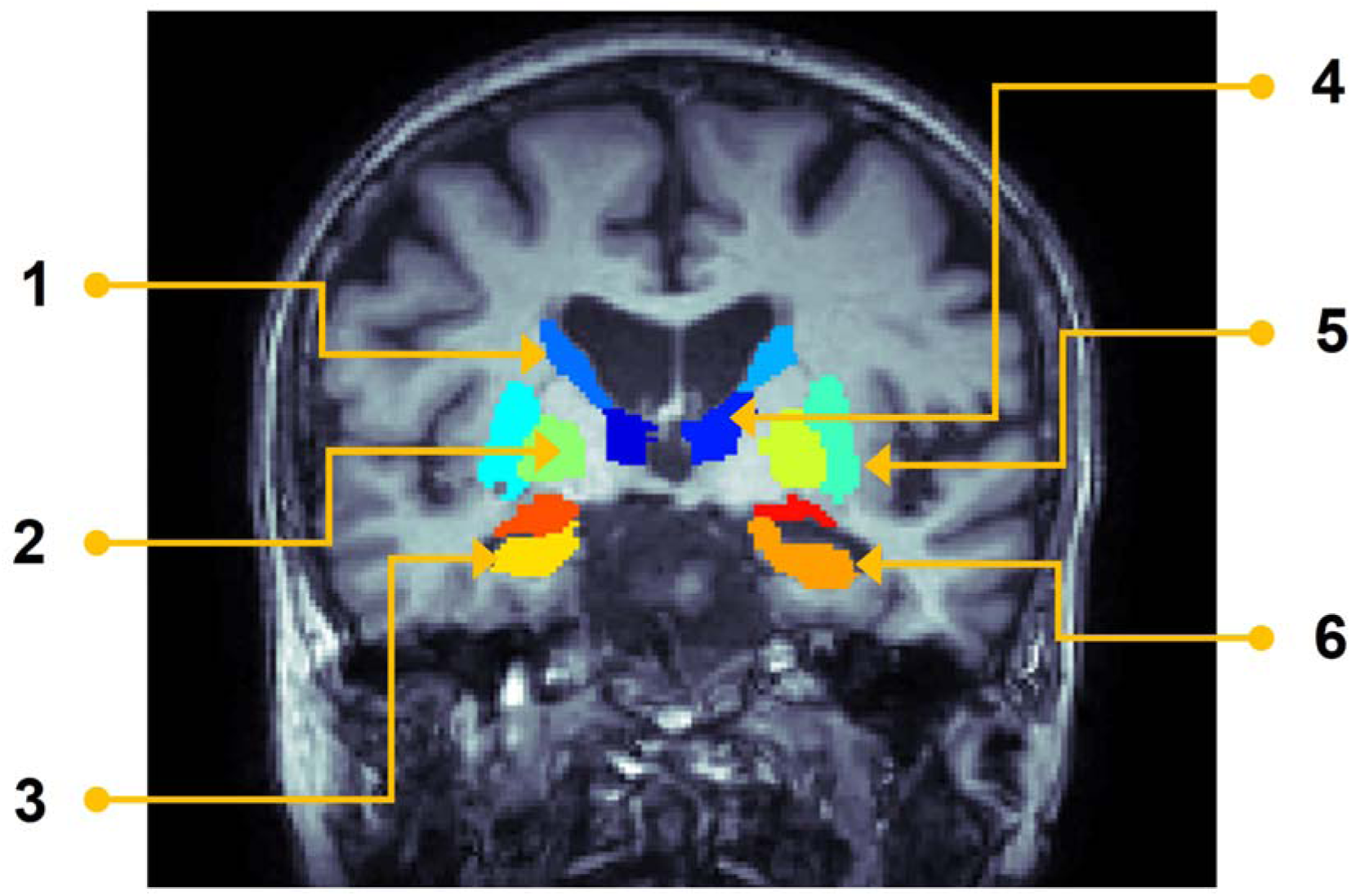
Frontal slice of structural MRI of a healthy subject with segmentation of 14 subcortical structures (7 structures bilaterally) according to the built-in atlas FreeSurfer 6.0.0. 1 – caudate nucleus; 2 – globus pallidus; 3 – amygdala; 4 – thalamus; 5 – putamen; 6 – hippocampus; Not shown – nucleus accumbens.

All MRI processing used standardized, automated pipelines with quality control performed by trained personnel blinded to clinical group.

### Statistical analysis

Between-group comparisons of mean cortical thickness in both hemispheres and volumes of regions of interest^1^ were performed using R (version 4.2.1). Pairwise analysis of covariance (ANCOVA) was conducted with age and sex as covariates. For volumetric measurements, the estimated total intracranial volume (eTIV) was included as a covariate. Neuroimaging measures served as the dependent variable, and group membership as the predictor.

Normality of distributions was assessed using the Shapiro–Wilk test (*stats* package v. 4.2.1), homogeneity of variances using Levene’s test (*car* package v. 3.1-0), and heteroscedasticity of model residuals using the White test (*ncvTest* function, *car* package v. 3.1-0). In cases where the assumptions for ANCOVA were violated, a nonparametric equivalent was applied using the *sm.ancova* function in the *sm* package (v. 2.2-5.6). The effect sizes for between-group differences were estimated using Cohen’s d.

The assumption of a linear relationship between the covariate age and the dependent variables was not tested separately, as large-scale studies from the ENIGMA consortium have previously demonstrated linear associations between age and cortical thickness ^24^, as well as between age and the subcortical structures examined in the present study ^25^, within the age range analyzed here.

Significance levels were corrected for multiple comparisons in each pairwise analysis, based on the number of tests performed within that specific analysis. Corrections were applied separately for 1) cortical thickness and subcortical structure volumes (16 tests) and 2) hippocampal subregions (24 tests) using the false discovery rate (FDR) method (q = 0.05) ^26^ via the *mt.rawp2adjp* function from the R *multtest* package (v. 2.52.0).

For the correlation analysis with baseline MoCA scores, only MRI measures that demonstrated significant between-group differences in any of the performed contrasts were included. This was performed in both the entire clinical group and in the outcome-based subgroups. Linear regression models were used, and all analyses were performed using R software (version 4.2.1). All variables were centered, scaled, and Box-Cox transformed with automatic lambda selection (*caret* package version 6.0-81). Age and estimated total intracranial volume (for volumetric measurements) were included in the models as additional covariates. Partial correlation coefficients were calculated using the formula 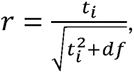 where t*_i_* is the t-statistic for the predictor of interest and *df* is the model degrees of freedom (*n*-*k*-1, where *n* is the number of observations and *k* is the number of predictors in the regression model) ^27^.

The significance levels of the regression coefficients of interest were corrected for multiple comparisons according to the number of correlations tested, using the false discovery rate (FDR) method (q = 0.05) ^26^ via the *mt.rawp2adjp* function from the R *multtest* package (v. 2.52.0). Corrections were performed separately for each subgroup and separately for 1) mean cortical thickness and subcortical structure volumes and 2) hippocampal subregion volumes.

Complete-case analysis was used to ensure there were no missing values in the primary neuroimaging or outcome variables.

## Results

### Comparison of Morphometric Measures Between the aMCI Group and Healthy Controls

Patients with aMCI as a whole demonstrated significantly smaller bilateral hippocampal volumes and volumes of most hippocampal subfields, as well as reduced left amygdala volume compared with healthy controls. No significant differences were found in the mean cortical thickness. Table 2 presents the results of these analyses.

**Table 2.** Results of Intergroup Comparisons between Patients with aMCI and Healthy Subjects.

| Region | aMCI (n=28)<br>Mean±SD | Control<br>(n=27)<br>Mean±SD | F | p-value | Cohen's<br>d | 95% CI |
| --- | --- | --- | --- | --- | --- | --- |
| **Left hemisphere** |  |  |  |  |  |  |
| **Mean cortical thickness** | 2.40±0.09 | 2.44±0.07 | 3.0 | .0913 | 0.5 | -0.1/1.0 |
| **Amygdala** | 1140±257 | 1309±218 | 8.8 | .0046 | 0.7 | 0.2/1.3 |
| **Hippocampus** | 2895±482 | 3219±351 | 12.5 | .0009 | 0.8 | 0.2/1.3 |
| **Hippocampal tail** | 429±82 | 485±65 | 10.3 | .0023 | 0.7 | 0.2/1.3 |
| **Subiculum** | 375±64 | 419±43 | 11.9 | .0011 | 0.8 | 0.2/1.4 |
| **CA1** | 537±99 | 594±84 | 7.1 | .0104 | 0.6 | 0.1/1.2 |
| **Presubiculum** | 274±51 | 302±35 | 6.8 | .0118 | 0.6 | 0.1/1.2 |
| **Molecular layer of hippocampus** | 480±91 | 535±65 | 9.5 | .0034 | 0.7 | 0.1/1.3 |
| **Granule cell layer of dentate gyrus** | 248±49 | 271±33 | 6.2 | .0162 | 0.5 | 0.0/1.1 |
| **CA3** | 175±34 | 191±30 | 4.8 | .033 | 0.5 | -0.1/1.0 |
| **CA4** | 216±40 | 237±28 | 7.3 | .0093 | 0.6 | 0.0/1.1 |
| **Fimbria** | 55±25 | 67±18 | 5.3 | .0258 | 0.6 | 0.0/1.1 |
| **Hippocampus-amygdala transition area** | 47±12 | 56±11 | 10.1 | .0026 | 0.8 | 0.2/1.4 |
| **Right hemisphere** |  |  |  |  |  |  |
| **Mean cortical thickness** | 2.40±0.09 | 2.44±0.08 | 3.5 | .0689 | 0.5 | -0.1/1.0 |
| **Hippocampus** | 2999±438 | 3273±350 | 10.9 | .0018 | 0.7 | 0.1/1.2 |
| **Subiculum** | 375±52 | 420±47 | 16.1 | .0002 | 0.9 | 0.4/1.5 |
| **CA1** | 563±85 | 609±72 | 6.5 | .0137 | 0.6 | 0.0/1.1 |
| **Presubiculum** | 256±39 | 281±35 | 9.9 | .0028 | 0.7 | 0.1/1.3 |
| **Molecular layer of hippocampus** | 493±78 | 545±62 | 11.8 | .0012 | 0.7 | 0.2/1.3 |
| **Granule cell layer of dentate gyrus** | 263±47 | 284±37 | 5.4 | .0243 | 0.5 | -0.1/1.0 |
| **CA3** | 194±39 | 211±32 | 5.0 | .0294 | 0.5 | -0.1/1.0 |
| **CA4** | 231±38 | 248±31 | 5.4 | .0237 | 0.5 | -0.1/1.0 |
| **Fimbria** | 55±21 | 68±15 | 10.6 | .0021 | 0.8 | 0.2/1.3 |
| **Hippocampus-amygdala transition area** | 52±10 | 60±9 | 13.4 | .0006 | 0.9 | 0.3/1.4 |

Note: Values are presented as means±standard deviations (mm<sup>3</sup> for volume, mm for cortical thickness). p-values after FDR correction. aMCI = amnesic mild cognitive impairment; aMCI-AD = converters to Alzheimer's disease dementia.

### Between-Group Comparisons of Clinical aMCI Subgroups with Different Outcomes and Healthy Controls

The stable aMCI subgroup showed smaller bilateral hippocampal volumes and volumes of several hippocampal subfields compared to healthy controls, while no differences in cortical thickness were observed (Table 3A, Figure 3A).

**Figure 3.**
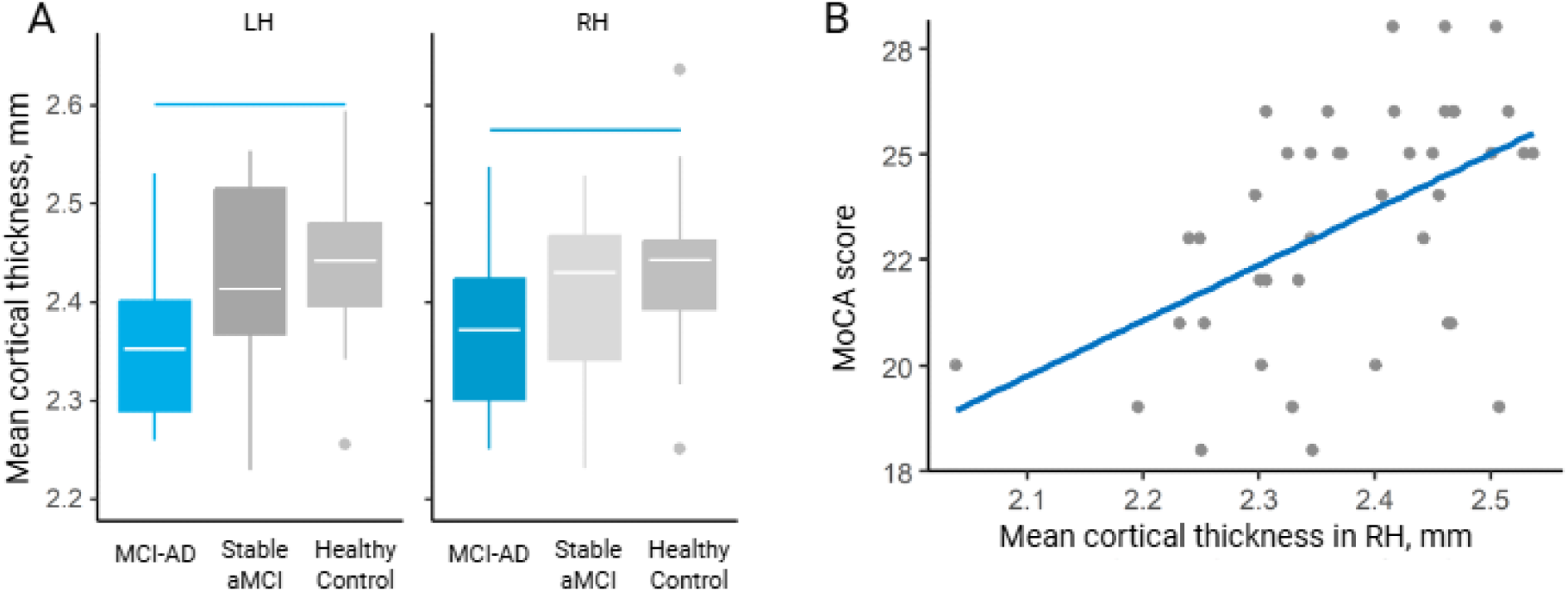
A: Boxplots of cortical thickness of the left (LH) and right hemispheres (RH) for clinical subgroups with different outcomes (MCI-AD is a group with dementia due to Alzheimer’s disease diagnosed during follow-up, Stable aMCI is a stable subgroup of patients with aMCI and healthy control group. B: Scatter plot: MoCA scores and cortical thickness in the right hemisphere of the total group of patients with aMCI (n = 41). R = 0.5, p = .0018.

**Table 3.** Results of Intergroup Comparisons between Patients with aMCI with Different Outcomes and Healthy Subjects.

**A. Stable aMCI Subgroup and Healthy Participants**
|  | **Stable aMCI**<br>**(n=17)**<br>**Mean±SD** | **Control**<br>**(n=24)**<br>**Mean±SD** | **F** | **p-**<br>**value** | **Cohen's**<br>**d** | **95% CI** |
| --- | --- | --- | --- | --- | --- | --- |
| **Age, years** | 71±10.8 | 70.7±8.2 | 0 | 0.9110 | 0 | -0.6/0.7 |
| **Left hemisphere** |  |  |  |  |  |  |
| **Mean cortical thickness** | 2.40±0.14 | 2.43±0.07 | 1.5 | .2352 | -0.3 | -1.0/0.3 |
| **Hippocampus** | 2970±416 | 3201±368 | 8.7 | .0056 | -0.6 | -1.2/0.1 |
| **Subiculum** | 383±57 | 417±45 | 8.8 | .0052 | -0.7 | -1.3/0.0 |
| **Molecular layer of hippocampus** | 494±74 | 533±69 | 7.2 | .0109 | -0.5 | -1.2/0.1 |
| **Hippocampus-amygdala transition area** | 48±13 | 57±11 | 6.9 | .0125 | -0.8 | -1.4/-0.1 |
| **Right hemisphere** |  |  |  |  |  |  |
| **Mean cortical thickness** | 2.38±0.13 | 2.43±0.08 | 3.6 | .0639 | -0.5 | -1.2/0.1 |
| **Hippocampus** | 3005±393 | 3255±356 | 9.7 | .0037 | -0.7 | -1.3/0.0 |
| **Subiculum** | 373±50 | 417±45 | 15.6 | .0004 | -1.0 | -1.6/-0.3 |
| **Presubiculum** | 251±37 | 280±35 | 10.1 | .0030 | -0.8 | -1.5/-0.1 |
| **Molecular layer of hippocampus** | 495±69 | 542±63 | 10.3 | .0028 | -0.7 | -1.4/0.0 |
| **Fimbria** | 56±22 | 68±14 | 6.7 | .0141 | -0.7 | -1.4/-0.1 |

Note: Values are presented as means±standard deviations (mm<sup>3</sup> for volume, mm for cortical thickness). p-values after FDR correction. aMCI = amnesic mild cognitive impairment.

**B. Group with Alzheimer's Disease Dementia Diagnosed at Follow-up and Healthy Study Participants**
|  | **aMCI-AD**<br>**Mean±SD** | **Control (n=24)**<br>**Mean±SD** | **F** | **p-value** | **Cohe**<br>**n's d** | **95% CI** |
| --- | --- | --- | --- | --- | --- | --- |
| **Age, years** | 74.4±7.6 | 70.7±8.2 | 2.1 | .1600 | 0.5 | -0.2/1.1 |
| **Left hemisphere** |  |  |  |  |  |  |
| **Mean cortical thickness** | 2.36±0.08 | 2.43±0.07 | 10.2 | .0029 | -1 | -1.7/-0.3 |
| **Amygdala** | 1056±191 | 1306±231 | 16.6 | .0002 | -1.2 | -1.9 /-0.5 |
| **Hippocampus** | 2623±460 | 3201±368 | 30.5 | <.0001 | -1.4 | -2.1/-0.7 |
| **Hippocampal tail** | 422±63 | 472±56 | 9.5 | .004 | -0.8 | -1.5/-0.2 |
| **Subiculum** | 331±69 | 417±45 | 29.9 | <.0001 | -1.6 | -2.3/-0.8 |
| **CA1** | 488±90 | 596±89 | 20.2 | .0001 | -1.2 | -1.9/-0.5 |
| **Presubiculum** | 253±54 | 299±35 | 14.7 | .0005 | -1.1 | -1.8/-0.4 |
| **Molecular layer of hippocampus** | 424±89 | 533±69 | 27.5 | <.0001 | -1.4 | -2.1/-0.7 |
| **Granule cell layer of dentate gyrus** | 217±47 | 270±35 | 24.3 | <.0001 | -1.3 | -2/-0.6 |
| **CA3** | 157±33 | 192±32 | 14.4 | .0006 | -1.1 | -1.8/-0.4 |
| **CA4** | 194±39 | 236±29 | 21 | .0001 | -1.2 | -1.9/-0.5 |
| **Fimbria** | 37±22 | 67±18 | 28.8 | <.0001 | -1.5 | -2.3/-0.8 |
| **Hippocampus-amygdala transition area** | 44±8 | 57±11 | 21.7 | <.0001 | -1.3 | -2/-0.6 |
| **Right hemisphere** |  |  |  |  |  |  |
| **Mean cortical thickness** | 2.36±0.09 | 2.43±0.08 | 7.9 | .0080 | -0.9 | -1.6/-0.2 |
| **Amygdala** | 1282±218 | 1490±244 | 11.2 | .002 | -0.9 | -1.6/-0.2 |
| **Nucleus accumbens** | 376±71 | 439±81 | 9.7 | .0037 | -0.8 | -1.5/-0.1 |
| **Hippocampus** | 2796±480 | 3255±356 | 19.7 | .0001 | -1.1 | -1.8/-0.4 |
| **Subiculum** | 347±56 | 417±45 | 30.2 | <.0001 | -1.4 | -2.2/-0.7 |
| **CA1** | 518±91 | 609±72 | 17.2 | .0002 | -1.1 | -1.8/-0.4 |
| **Presubiculum** | 242±41 | 280±35 | 18.1 | .0002 | -1 | -1.7/-0.3 |
| **Molecular layer of hippocampus** | 450±84 | 542±63 | 24.5 | <.0001 | -1.3 | -2/-0.6 |
| **Granule cell layer of dentate gyrus** | 236±49 | 283±40 | 17.1 | .0002 | -1.1 | -1.8/-0.4 |
| **CA3** | 178±38 | 210±34 | 10.9 | .0022 | -0.9 | -1.6/-0.2 |
| **CA4** | 212±41 | 247±33 | 13.3 | .0009 | -0.9 | -1.6/-0.3 |
| **Fimbria** | 45±25 | 68±14 | 20.2 | .0001 | -1.2 | -1.9/-0.5 |
| **Hippocampus-amygdala transition area** | 48±6 | 60±9 | 26.5 | <.0001 | -1.5 | -2.2/-0.7 |

Note: Values are presented as means±standard deviations (mm<sup>3</sup> for volume, mm for cortical thickness). p-values after FDR correction. aMCI = amnesic mild cognitive impairment.

C. Stable aMCI Subgroup and Group with AD Dementia Diagnosed at Follow-up
|  | Stable<br>aMCI<br>Mean±SD | aMCI-AD<br>Mean±SD | F | p-value | Cohen's<br>d | 95% CI |
| --- | --- | --- | --- | --- | --- | --- |
| Age, years | 71±10.8 | 74.4±7.6 | 1.1 | .3120 | -0.4 | -1.1/0.4 |
| Left hemisphere |  |  |  |  |  |  |
| Mean cortical thickness | 2.4±0.14 | 2.36±0.08 | 1.3 | .2675 | 0.4 | -0.3/1.1 |
| Subiculum | 383±57 | 331±69 | 7.7 | .0096 | 0.8 | 0.1/1.6 |
| CA1 | 568±83 | 488±90 | 8.1 | .0082 | 0.9 | 0.2/1.7 |
| Molecular layer of<br>hippocampus | 494±74 | 424±89 | 8.3 | .0075 | 0.9 | 0.1/1.6 |
| Granule cell layer of<br>dentate gyrus | 256±44 | 217±47 | 10.<br>7 | .0028 | 0.9 | 0.1/1.6 |
| CA3 | 181±33 | 157±33 | 8.5 | .0069 | 0.7 | 0/1.5 |
| CA4 | 221±35 | 194±39 | 7.8 | .0093 | 0.7 | 0/1.5 |
| Right hemisphere |  |  |  |  |  |  |
| Mean cortical thickness | 2.38±0.13 | 2.36±0.09 | 0.3 | .5799 | 0.2 | -0.5/0.9 |
| Granule cell layer of<br>dentate gyrus | 268±42 | 236±49 | 6.8 | .0144 | 0.7 | 0/1.4 |
| Hippocampus-amygdala transition<br>area | 55±11 | 48±6 | 5.4 | .0278 | 0.8 | 0/1.5 |

Note: Values are presented as means±standard deviations (mm<sup>3</sup> for volume, mm for cortical thickness). p-values after FDR correction. aMCI = amnesic mild cognitive impairment.

Patients who were diagnosed with AD dementia during the longitudinal follow-up demonstrated bilateral cortical thinning, as well as reduced volumes of the amygdala, hippocampus, and most hippocampal subfields compared with healthy controls (Table 3B, Figure 3A).

The two aMCI subgroups with different clinical outcomes differed only in the volumes of several hippocampal subfields (Table 3C).

### Correlation Analysis

The only result that survived correction for multiple comparisons was a positive correlation between cortical thickness in the right hemisphere and the baseline MoCA score in the total sample of patients with aMCI (p = .0018, r = 0.50) (Figure 3B).

## Discussion

In the present study, we aimed to identify structural brain features associated with aMCI and to determine whether baseline morphometric measures could differentiate patients who subsequently progress to AD dementia from those who remain stable. We hypothesized that aMCI would be characterized by hippocampal subfields’ volumes reduction, with more pronounced and widespread cortical and subcortical involvement in individuals with progressive dementia.

The present study revealed reduced volumes of the hippocampus and several of its subregions both in the overall aMCI group and in both outcome-based subgroups. These findings are consistent with data from other studies ^5,9,28,29^ and indicate a distributed structural deficit in the hippocampal complex in MCI. Simultaneously, the hippocampal fissure and parasubiculum appeared to be relatively preserved in aMCI. Similar results have been reported by Xiao et al. ^30^ The preservation of the parasubiculum in patients with MCI is also supported by findings from Russian authors ^9^. Moreover, we did not observe reduced volumes of the parasubiculum even in the subgroup with the most pronounced structural abnormalities, patients who progressed to AD dementia during follow-up. This is consistent with reports of parasubiculum preservation in certain subgroups of patients already diagnosed with AD ^31^.

The next subcortical structure showing reduced volume in the overall clinical group and in the aMCI-AD subgroup was the amygdala. Notably, volume reduction was observed in the left, but not the right, amygdala in the total aMCI group, which aligns with the results of a meta-analysis by Zhang et al. ^5^ The left amygdala is involved in the voluntary cognitive control of emotional processes and verbal processing of emotional information ^32^. The involvement of the amygdala in the pathological process is particularly noteworthy given the growing interest in behavioral symptoms, such as mild anxiety and depressive symptoms, in patients with MCI ^33^. Furthermore, knowledge of the functional specialization of the left amygdala (the aforementioned voluntary control and verbal processing of emotional information) may, in theory, be considered in psychotherapeutic interventions for patients with MCI.

At the same time, we did not find a statistically significant reduction in left amygdala volume in the stable aMCI subgroup (1199±242 vs 1306±231, F = 3.3, p =.0785, Cohen’s d = −0.5, 95% CI: −1.1; 0.2). This may be attributable to both insufficient statistical power due to the limited sample size and the relatively more preserved overall brain structural state in this subgroup.

Regarding the other subcortical structures, we found no significant volume differences in either the aMCI group or the two clinical subgroups. This finding is consistent with data from recent studies ^34^, including the latest available meta-analysis ^5^, which also reported abnormalities primarily in the hippocampus and amygdala, but not in other subcortical structures.

Another important finding was the bilateral reduction in the mean cortical thickness observed in patients with aMCI who were subsequently diagnosed with AD dementia. This reduction was not observed in the stable aMCI subgroup or the aMCI group in total (Figure 3A). Together with the positive correlation between MoCA scores and cortical thickness (Figure 3B), these results allowed us to cautiously consider the degree of cortical gray matter deficit as a potentially meaningful neuroimaging prognostic marker. Moreover, this finding aligns with the review by Drago et al., in which the severity of global cerebral cortical atrophy was classified among the markers with a high degree of validity ^13^.

When assessing the associations between the structural measures examined and MoCA scores in the entire clinical sample (n = 41), a positive correlation was found between the mean cortical thickness in the right hemisphere and the MoCA score. This moderate correlation suggests that greater right-hemisphere cortical thickness in patients with aMCI may be associated with better cognitive functioning at the time of the assessment. This result can be interpreted in the context of altered lateralization in patients with MCI. It has been proposed that the greater involvement of the right hemisphere in cognitive activity may serve as a compensatory mechanism ^35^. In this regard, it is also noteworthy that a somewhat smaller number of structures in the right hemisphere were involved in the atrophic process, specifically, the absence of significant differences between the aMCI group and healthy controls in the volumes of the hippocampal tail and amygdala. Thus, it can be cautiously hypothesized that there is hemispheric asymmetry of pathological changes in aMCI, characterized by greater involvement of the left hemisphere in the pathogenetic process, while right-hemispheric structures and their functional reserve may contribute to the development of compensatory mechanisms.

### Limitations

This study has several limitations. First, the sample size was relatively small, especially for defined clinical subgroups. This limited statistical power may have prevented the detection of smaller but potentially meaningful differences between subgroups and reduced the precision of the effect size estimates. Second, the diagnosis of aMCI and subsequent diagnosis of AD dementia were made using ICD-10-based clinical criteria and cognitive assessment, without the use of cerebrospinal fluid or amyloid/tau PET imaging. Consequently, the possibility of including patients with aMCI due to non□AD pathologies could not be excluded. Third, despite careful age□ and sex□matching for the main comparisons, the original overall groups differed significantly in age, and the matched control sample (n□=□27) only partially overlapped with the subgroups; residual confounding by age or other unmeasured factors could not be ruled out. Fourth, the follow□up interval was relatively short, and the subgroup with cognitive decline that did not meet the criteria for the diagnosis of dementia in AD (n□=□5) was too small for meaningful analysis, precluding a more granular view of the progression trajectory. Fifth, structural MRI was acquired at a single time point; longitudinal imaging is needed to directly assess within□subject atrophy rates, which are known to be strong predictors of conversion. Finally, the findings were obtained from a single centre and lacked external validation in an independent cohort, thus limiting generalizability. Future studies with larger multicenter samples, biomarker□confirmed AD pathology, and longitudinal imaging are warranted to validate the proposed prognostic markers.

## Conclusions

The present study revealed structural brain abnormalities in patients with aMCI, including bilateral reductions in hippocampal and hippocampal subfield volumes as well as reduced amygdala volume (particularly on the left), with relative preservation of other subcortical structures.

Smaller bilateral cortical gray matter thickness may be considered a potentially significant prognostic marker for conversion to AD dementia in patients with aMCI. However, this assumption requires further verification and will be the subject of future studies.

## Data Availability

All data produced in the present study are available upon reasonable request to the authors

## Acknowledgement

We sincerely thank all study participants and their study partners.

## Statements and Declarations

## Ethical considerations

This non-interventional observational study was performed in accordance with the principles of the Helsinki Declaration (1964) and its later amendments (1975–2013) and was approved by the Local Ethics Committee of the Federal State Budgetary Scientific Institution “Russian Mental Health Research Centre” (Protocol No. 920 25-Dec-2023).

## Consent to participate

All patients provided written informed consent to participate in the observational program.

## Consent for publication

All images were acquired after participants had provided informed consent for publication.

## Data availability

The data that support the findings of this study are available from the corresponding author, N. Cherkasov, upon reasonable request.

## Footnotes

1 Thalamus, caudate nucleus, putamen, globus pallidus, amygdala, nucleus accumbens, hippocampus; hippocampal subregions: hippocampal tail, subiculum, CA1, hippocampal fissure, presubiculum, parasubiculum, molecular layer of the hippocampus, granule cell layer of the dentate gyrus, CA3, CA4, fimbria, and hippocampus-amygdala transition area.

